# When “Shelter-in-Place” Isn’t Shelter That’s Safe: A Rapid Analysis of Domestic Violence Case Differences During the COVID-19 Pandemic and Stay-at-Home Orders

**DOI:** 10.1101/2020.05.29.20117366

**Authors:** Molly M. McLay

## Abstract

**Purpose:** This study explored the COVID-19 pandemic’s impacts on domestic violence (DV) with the following research questions: 1) Did DV occurring during the pandemic differ on certain variables from cases occurring on a typical day the previous year? 2) Did DV occurring after the implementation of shelter-in-place orders differ (on these same variables) from cases occurring prior to shelter-in-place orders?

**Methods:** Two logistic regression models were developed to predict DV case differences before and during the pandemic. DV reports (N=4618) were collected from the Chicago Police Department. Cases from March 2019 and March 2020 were analyzed based on multiple variables. One model was set to predict case differences since the pandemic began, and another model was set to predict case differences during the shelter-in-place period later that month.

**Results:** Both models were significant with multiple significant predictors. During the pandemic period, cases with arrests were 20% less likely to have occurred, and cases at residential locations were 22% more likely to have occurred. During the shelter-in-place period, cases at residential locations were 64% more likely to have occurred, and cases with child victims were 67% less likely to have occurred.

**Conclusions:** This study offers a rapid analysis of DV case differences since the pandemic and shelter-in-place began. Additional variables and data sources could improve model explanatory power. Research, policy, and practice in this area must pivot to focus on protecting children whose access to mandated reporters has decreased and moving victims out of dangerous living situations into safe spaces.

## Background

The COVID-19 pandemic has wreaked havoc across the world since late 2019, causing nearly 5 million individuals to fall ill and over 300,000 to die, according to the Johns Hopkins Coronavirus Resource Center as of mid-May 2020 (Dong, Du, & Gardner, 2020). COVID-19’s impact on the United States (U.S.) began in late January 2020, and the numbers of individuals impacted is still rising exponentially, with a total of 1,508,291 cases and 90,340 deaths (Dong, Du, & Gardner, 2020). While research has jump-started to develop testing, treatment, and vaccines, the impact of the COVID-19 on social factors cannot be ignored. The Centers for Disease Control and Prevention (CDC) have recommended that individuals stay six feet away apart, not gather in groups, and only venture out for necessities (CDC, 2020b). As a result, businesses closed or reduced operations, schools moved online, and many individuals began working from home (CDC, 2020a). In March 2020, a number of states began issuing statewide stay-at-home or “shelter-in-place” orders further restricting their residents, including Illinois (Petrella, St. Clair, Johnson, & Pratt, 2020), which, at the time of this study, had the third largest number of COVID-19 cases of all states (Dong, Du, & Gardner, 2020). Given the unique circumstances of the COVID-19 pandemic, research is needed to identify its impacts on individuals, families, and communities.

With social distancing recommendations and shelter-in-place orders making abrupt changes to the ways people interact, this study aimed to understand the pandemic’s potential impacts on one volatile form of interpersonal interaction: domestic violence. Utilizing the U.S. Department of Justice (DOJ) definition, domestic violence includes violence toward current or former intimate partners, family members (including children), or anyone with whom the perpetrator shares a home (U.S. DOJ, 2019). Such violence has far-reaching impacts; 1 in 3 women and 1 in 4 men in the U.S. have experienced some form of violence from an intimate partner over the life course (Black et. al., 2011), and 68-80% of children have been exposed to violence in the home (Jaffe, 2018). Domestic violence has devastating effects on its victims and their loved ones, with mental health consequences and physical injuries being common; Black and colleagues (2011) noted that women with a history of violence were nearly three times more likely to rate their physical or mental health as poor, which may be compounded in the midst of a global pandemic. Domestic violence can also be fatal; approximately 55% of female homicide victims are killed by an intimate partner (CDC, 2019).

Research on domestic violence prevention and treatment during the pandemic is ongoing. At the time of this study, scholars had published commentaries regarding COVID-19’s potential impact on mental health issues broadly (Kumar & Navar, 2020) and domestic violence more specifically (Abramson, 2020; Bosman, 2020; Usher, Bhullar, Durkin, Gyamfi, & Jackson, 2020; Van Gelder et. al., 2020). However, at that time, no known peer-reviewed studies had analyzed domestic violence rates in light of the pandemic. With more individuals at home due to shelter-in-place orders, violence may happen more frequently behind closed doors and in increased isolation. Individuals may also have decreased access to resources, given that especially given that first responders, service providers, and mandated reporters may experience reduced staffing and increased virtual interactions (CDC, 2020a).

These changes to access may be felt more acutely by individuals in communities with strict stay-at-home orders, like in Illinois, where schools were closed the end of the spring semester (Petrella, St. Clair, Johnson, & Pratt, 2020). During this time of increased stress, financial strain, and fear for many (Brooks et. al., 2020), changes may then be seen in domestic violence, the factors comprising it, and the individuals experiencing it. Research is needed to understand what domestic violence looks like in this unprecedented moment, so that prevention and treatment can shift in order to appropriately respond.

## Research Questions

This study analyzed the potential impacts of the COVID-19 pandemic on domestic violence reports in the U.S. through logistic regression analysis. Two models were developed to predict changes in domestic violence before and during the pandemic at different time points, utilizing data from a large Illinois city before and during the height of key policy changes. This study analyzed relationships between the timeframe of domestic violence occurrence and a number of key variables. This study poses the following research questions:

1. Will domestic violence occurring during the COVID-19 pandemic in March 2020 differ on certain variables (the presence of a sex offense, weapon use, resulting arrest, presence of child victims, and residential locationality) from domestic violence occurring on a typical day in March 2019?
2. Will domestic violence occurring after the implementation of COVID-19 shelter-in-place orders on March 21, 2020, differ on these same variables from domestic violence occurring during the pandemic but prior to shelter-in-place orders?

## Methods

### Sampling

The study’s sample is comprised of police reports from the Chicago Police Department (CPD). Data was downloaded from the Chicago Data Portal in early May 2020. Reports on cases that occurred during the months of March 2019 and March 2020 were utilized. March 2019 was used as a “usual circumstances” comparison to the March 2020 calls, which occurred as the COVID-19 pandemic took hold on this city. March 2020 is also the month in which Chicago’s first shelter-in-place order was put into effect (Petrella, St. Clair, Johnson, & Pratt, 2020), which became a point of analysis.

There were 19,747 police reports documented by CPD for March 2019 cases, and there were 15,852 reports for March 2020 cases. The data was narrowed to any reports considered domestic incidents, for a total of 6,559 reports, and subsequently imported from a CSV file into a Stata data file. The sample was then reduced only to reports involving some kind of physical or sexual violence toward a person. These offenses included any kind of assault, any kind of battery, homicide, criminal sexual assault, any sex offense against an adult, and any physical or sexual offenses against children. (Arson, damage to property, trespassing, “deceptive practices” such as identity theft, robbery, theft, and weapons violations were excluded. Stalking, kidnapping, verbal harassment, and violations of orders of protection were also not included, as these could not be confirmed to have a component of physical or sexual violence.) The final sample consisted of 4,618 police reports. This dataset did not include any demographic information about victims or perpetrators; information was limited to basic case descriptors, crime code and characteristics, location data, and basic date/time information.

### Variables and Instrumentation

Two dependent variables were analyzed: occurrence of domestic violence during the pandemic or not, and occurrence of domestic violence during shelter-in-place order or not. These two dependent variables, as well as several independent variables, are described as follows:

### Occurrence of domestic violence during COVID-19 pandemic

One dependent variable for this study was whether or not a domestic violence case occurred during the COVID-19 pandemic. For this study, March 2019 and March 2020 police calls were the two groups analyzed, and these represented the dummy coding for this variable, with March 2020 cases being the group of interest and March 2019 being the reference group. This coding reflected the study’s purpose of determining any significant differences in the composition of domestic violence cases before and during the pandemic. To create this variable, any report for domestic violence occurring between March 1-31, 2020, was coded as “pandemic” = “1.” Any report for domestic violence occurring between March 1-31, 2019, was coded as “pandemic” = “0.”

### Occurrence of domestic violence during shelter-in-place order

The other dependent variable for this study was whether or not a domestic violence case occurring during the pandemic happened before or after Chicago’s shelter-in-place order took effect, as this order placed additional constraints on individual’s interactions and whereabouts. The shelter-in-place or “stay-at-home” order went into effect on March 21, 2020, at 5:00pm (Petrella, St. Clair, Johnson, & Pratt, 2020). Cases occurring on or after this time comprised the group of interest, and cases between March 1, 2020, and the start time of the shelter-in-place order on March 21, 2020, comprised the reference group. This coding reflected the study’s purpose of determining any significant differences in the composition of domestic violence cases before and during shelter-in-place. To create this variable, any report for domestic violence occurring between March 21 at 5:00pm and March 31 at 11:59pm, was coded as “stayhome” = “1.” Any report for domestic violence occurring between March 1 at 12:00am and March 21 at 4:59pm was coded as “stayhome” = “0.”

### Presence of sex crime

The presence of a sex crime during domestic violence was one independent variable analyzed. For the purposes of this research, sex crime was operationalized as any of the following crimes: criminal sexual assault, sex offense, and any sexual offenses against children. A variable called “sexcrime” was created. Any case containing one of these elements was coded as “1,” and any case that did not was coded as “0.”

### Use of weapon

Use of a weapon during domestic violence was another independent variable contributing to these analyses. In CPD records, all cases involving weapons were designated as “aggravated” per the criminal code. A variable called “weapon” was created. Any case involving the use of a weapon was coded as “1,” and any case that did not was coded as “0.”

### Whether or not arrest was made

The issuance of an arrest during a domestic violence report was another independent variable contributing to these analyses. In CPD records, all cases in which an arrest was made were coded as such. A variable called “arrest” was created. Any case in which an arrest was made was coded as “1,” and any case without was coded as “0.”

### Presence of child victim(s)

The presence of one or more child victims in a domestic violence report was another independent variable contributing to these analyses. In CPD records, cases involving children were categorized as “offenses against children.” A few crimes (n=7) in which child victims were referenced in descriptions, but not categorized under “offenses against children,” were manually recoded under this category. A variable called “child” was created, and any case involving a child victim was coded as “1.” Any cases without a reported child victim were coded as “0.” An interaction term was also created between presence of child victims and presence of sex crimes.

### Location of offense

A final independent variable was location—more specifically, whether or not the violence took place at someone’s residence. For the purposes of this research, residence was operationalized as any of the following locations: apartment; residence; nursing home facility; or the grounds, parking lot, or yard of any of these locations. Hotels were not included as residences, nor were any commercial locations or outside spaces unattached to a place where someone lived. A variable called “residence” was located, and any case taking place at one of these residences was coded as “1.” All cases taking place elsewhere were coded as “0.”

### Data Analysis Plan and Rationale

Logistic regression analysis was chosen as the primary data analysis tool for this study, as a number of independent variables were of interest, thus justifying the need for a multivariate analysis. Utilizing a maximum likelihood estimator, logistic regression analysis selects coefficients that maximize the probability of reproducing the sample data. Since two “whether or not” situations were being examined, it makes sense to analyze them in terms of probabilities or likelihood of each event occurring. Two models with dichotomous dependent variables were developed: one predicted whether or not a domestic violence case took place during the COVID-19 pandemic, and the other predicted whether or not a domestic violence case took place after the shelter-in-place order went into effect. The size of this sample was much greater than the minimum needed for logistic regression (anywhere from 100-500 observations; Long, 1997). In addition to ensuring independence of observations and proper sample size, multicollinearity and influential data checks were performed to ensure these assumptions of logistic regression were met. Each model utilized in the study and their formal specifications are outlined as follows.

### Model 1: Occurrence of domestic violence during COVID-19, regressed on multiple variables

The first logistic regression model featured occurrence of domestic violence before or during the COVID-19 pandemic regressed on the following independent variables: presence of a sex crime, weapon use, resulting arrest, presence of child victim(s), and residential locationality. Under this model, it is expected that, all other things being equal: when a sex crime is present, probability of domestic violence occurring during the COVID-19 pandemic changes; when a weapon is used, probability of domestic violence occurring during this pandemic changes; when an arrest is made, probability of domestic violence occurring during this pandemic changes; when a child victim is present, probability of domestic violence occurring during this pandemic changes; and when the location of the violence is a residence, probability of domestic violence occurring during this pandemic changes. The intercept in this model is the predicted log odds of domestic violence’s occurrence before or during the COVID-19 pandemic when there is no sex crime present, no weapon use, no resulting arrest, no child victim(s), and the location of the violence was not a residence. The formal model specification is as follows:

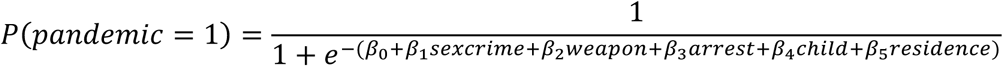

Two-tailed significance testing was performed on each regression coefficient, and odds ratios and their confidence intervals were reported. Two-tailed testing was chosen due to the unprecedented nature of the COVID-19 pandemic and its yet unknown effects on domestic violence. Given the exploratory nature of this study, this test is appropriate. Joint hypothesis testing and interaction effects were explored based on various bivariate analyses.

### Model 2: Occurrence of domestic violence during shelter-in-place order, regressed on multiple variables

The second logistic regression model featured occurrence of domestic violence before or during the shelter-in-place order regressed on the same independent variables as the first model: presence of a sex crime, weapon use, resulting arrest, presence of child victim(s), and residential locationality. Under this model, it is expected that, all other things being equal: when a sex crime is present, probability of domestic violence occurring during shelter-in-place changes; when a weapon is used, probability of domestic violence occurring during shelter-in-place changes; when an arrest is made, probability of domestic violence occurring during shelter-in-place changes; when a child victim is present, probability of domestic violence occurring during shelter-in-place changes; and when the location of the violence is a residence, probability of domestic violence occurring during shelter-in-place changes. The intercept in this model is the predicted log odds of domestic violence’s occurrence before or during the shelter-in-place order when there is no sex crime present, no weapon use, no resulting arrest, no child victim(s), and the location of the violence was not a residence. The formal model specification is below:

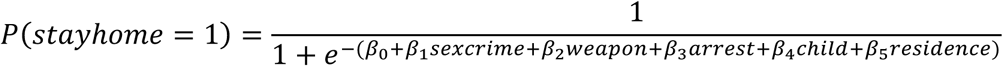

Similar to the previous model, two-tailed significance testing was performed on each regression coefficient, with odds ratios and confidence intervals reported. Two-tailed testing was chosen because of the unprecedented nature of the shelter-in-place orders and their yet unknown effects on domestic violence. Given the exploratory nature of this study, this test is appropriate. Similar to the first model, joint hypothesis testing using the likelihood ratio test examined whether or not weapon use and resulting arrest jointly contribute to the model fit. Also similarly, joint hypothesis testing and interaction effects were explored based on various bivariate analyses.

## Results

### Descriptive Statistics

The study’s dataset of March 2019 and March 2020 domestic violence cases contained 4,618 records, with complete information on sex offenses, weapon use, arrests made, child victims, and crime locations. At this size, the sample was large enough to meet the assumptions of logistic regression (Long, 1997). All variables were dichotomous, and independence of observations was met. Descriptive statistics are reported for March 2019 and March 2020 separately and in aggregate (Table 1).

**Table 1.**
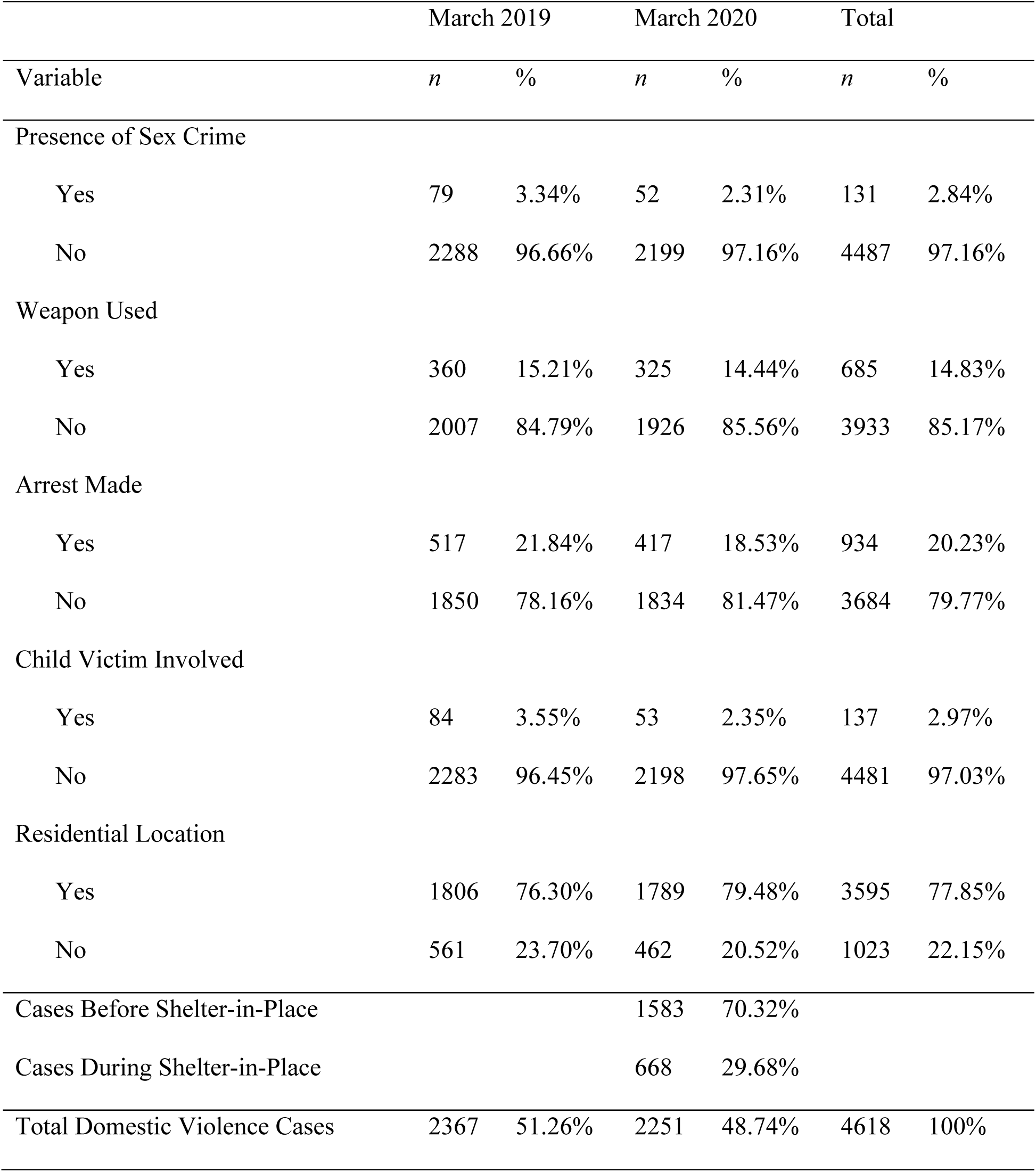
Descriptive Statistics of Domestic Violence Cases, March 2019 and March 2020, Chicago, IL

|  | March 2019 |  | March 2020 |  | Total |  |
| --- | --- | --- | --- | --- | --- | --- |
| Variable | *n* | % | *n* | % | *n* | % |
| Presence of Sex Crime |  |  |  |  |  |  |
| Yes | 79 | 3.34% | 52 | 2.31% | 131 | 2.84% |
| No | 2288 | 96.66% | 2199 | 97.16% | 4487 | 97.16% |
| Weapon Used |  |  |  |  |  |  |
| Yes | 360 | 15.21% | 325 | 14.44% | 685 | 14.83% |
| No | 2007 | 84.79% | 1926 | 85.56% | 3933 | 85.17% |
| Arrest Made |  |  |  |  |  |  |
| Yes | 517 | 21.84% | 417 | 18.53% | 934 | 20.23% |
| No | 1850 | 78.16% | 1834 | 81.47% | 3684 | 79.77% |
| Child Victim Involved |  |  |  |  |  |  |
| Yes | 84 | 3.55% | 53 | 2.35% | 137 | 2.97% |
| No | 2283 | 96.45% | 2198 | 97.65% | 4481 | 97.03% |
| Residential Location |  |  |  |  |  |  |
| Yes | 1806 | 76.30% | 1789 | 79.48% | 3595 | 77.85% |
| No | 561 | 23.70% | 462 | 20.52% | 1023 | 22.15% |
| Cases Before Shelter-in-Place |  |  | 1583 | 70.32% |  |  |
| Cases During Shelter-in-Place |  |  | 668 | 29.68% |  |  |
| Total Domestic Violence Cases | 2367 | 51.26% | 2251 | 48.74% | 4618 | 100% |

In terms of total domestic violence cases, 51.26% (n=2367) of the sample’s cases occurred in March 2019 and 48.74% (n=2251) took place in March 2020, meaning that there were slightly fewer cases during the pandemic month than there were during the previous year’s “typical” month. During March 2020, 70.32% (n=1583) of cases took place prior to the shelter-in-place, and 29.68% (n=668) of cases took place after. This finding is consistent with the fact that the shelter-in-place began on March 21—just over two-thirds of the way through the month.

In terms of overall trends, the overall sample contained cases with sex crimes 2.84% of the time, weapon use 14.83% of the time, arrests made 20.23% of the time, child victims involved 2.97% of the time, and residential locations involved 77.85% of the time. The amount that these varied by occurrence before or during the COVID-19 pandemic are described in Table 1 and was explored in the multiple logistic regression analyses.

### Bivariate Analyses

Bivariate analyses were conducted on a few independent variables of interest to determine potential need for interaction terms and joint hypothesis testing.

Several interaction effects were tested preliminarily. The most promising was the potential interaction between the presence of a sex crime and the presence of child victims, for both data-driven and theoretical reasons. Victims of sex crimes and child victims are two particularly vulnerable groups whose need for in-person mandated reporting and emergency medical resources may be particularly impacted during a pandemic and/or shelter-in-place period, and the experience of their intersection—child sexual victimization—is prevalent (Finkelhor, Shattuck, Turner, & Hamby, 2014). Chi-square analysis revealed a strong association between presence of a sex crime and presence of child victims (Table 2).

**Table 2.** Chi-Square Associations Between Independent Variables

| Sex Crime | Presence of Child Victim |  | Weapon Used | Arrest Made |  |
| --- | --- | --- | --- | --- | --- |
|  | No | Yes |  | No | Yes |
| No | 4406 | 81 | No | 3165 | 768 |
| Yes | 75 | 56 | Yes | 519 | 166 |
| *Note.* $\chi^2(1, N=4618)=741.21, p=.000$ | | | *Note.* $\chi^2(1, N=4618)=8.0094, p=.005$ | | |

Additionally, weapon use and the involvement of an arrest in domestic violence posed an interesting relationship. Previous research has pointed to a relationship between weapon use in domestic violence and subsequent arrest in that situation (Sorenson, 2017). Chi-square analysis revealed a significant association between these two variables (Table 2). Joint hypothesis testing using likelihood ratio was utilized on these two significantly associated variables, because the size and significance of their association was smaller than the association between sex crimes and child victims. Other bivariate relationships were analyzed, but they did not yield statistically significant results, and thus interactions between those variables were not built into final models.

### Logistic Regression Analyses

Models were constructed for both dependent variables after several experimental runs. Since chi-square analysis revealed a strong association between presence of a sex crime and presence of child victims, an interaction term of these two variables was included in preliminary runs. The interaction term was not significant for either model, so this term was dropped. Results of each model are presented in Tables 3 and 4 and are detailed as follows.

**Table 3.** Occurrence of Domestic Violence During COVID-19, Regressed on Multiple Variables

| Predictor Variable | Odds Ratio | Sign? | Coef. | Std. E | z | 95% CI OR |
| --- | --- | --- | --- | --- | --- | --- |
| Presence of Sex Crime | .753 | ? | -.284 | .149 | -1.43 | [.510, 1.11] |
| Use of Weapon | .972 | ? | -.028 | .081 | -.34 | [.825, 1.15] |
| Presence of Child Victim | .702 | ? | -.354 | .136 | -1.83 | [.480, 1.03] |
| Arrest Made | .804** | ? | -.218 | .059 | -2.95 | [.696, .930] |
| Residential Location | 1.224** | ? | .202 | .087 | 2.83 | [1.06, 1.41] |
| _cons | .868* |  | -.142 | .057 | -2.16 | [.763, .987] |
| Pseudo R<sup>2</sup> | .004 |  |  |  |  |  |
| Model Likelihood Ratio X<sup>2</sup> (df) | 24.12(5)*** |  |  |  |  |  |
| N | 4618 |  |  |  |  |  |

Note: \* p<.05, \*\* p<.01, \*\*\* p<.001, from two-tailed test.

**Table 4.** Occurrence of Domestic Violence During Shelter-in-Place, Regressed on Multiple Variables

| Predictor Variable | Odds Ratio | Sign? | Coef. | Std. E | z | 95% CI OR |
| --- | --- | --- | --- | --- | --- | --- |
| Presence of Sex Crime | 1.043 | ? | .042 | .358 | .12 | [.532, 2.05] |
| Use of Weapon | .835 | ? | -.180 | .114 | -1.33 | [.639, 1.09] |
| Presence of Child Victim | .329** | ? | -1.11 | .139 | -2.62 | [.144, .755] |
| Arrest Made | .960 | ? | -.041 | .115 | -.34 | [.759, 1.22] |
| Residential Location | 1.638*** | ? | .123 | .202 | 4.01 | [1.29, 2.09] |
| _cons | .028*** |  | .115 | .034 | -10.54 | [.238, .373] |
| Pseudo R<sup>2</sup> | .01 |  |  |  |  |  |
| Model Likelihood Ratio X<sup>2</sup> (df) | 27.08(5)** |  |  |  |  |  |
| N | 2251 |  |  |  |  |  |

Note: \* p<.05, \*\* p<.01, \*\*\* p<.001, from two-tailed test.

### Model 1: Occurrence of domestic violence during COVID-19, regressed on multiple variables

The results of the first logistic regression model, predicting occurrence of domestic violence during COVID-19 regressed on several key variables, provided interesting findings with low explanatory value per its pseudo R^2^ (Table 3).

In terms of overall model appropriateness, Hosmer-Lemeshow goodness-of-fit test indicated that the logistic response function was appropriate with a non-significant chi-square value of 9.07, p=.97. To check for multicollinearity problems, a variance inflation factor (VIF) test was run, resulting in a low mean VIF of 1.09, indicating no problems. Influential data was assessed with Cook’s distance and was not a problem for this model.

A chi-square test was then performed on the log-likelihood ratio, resulting in a statistically significant overall model (X^2^=24.12, df=5, p<.001). In addition to the significant intercept, the variables of resulting arrest and residential location were both significant. Odds ratios were then interpreted for arrest (.804) and residential location (1.224). All other things being equal, a domestic violence case with an arrest made was approximately 20% less likely (than cases without arrests) to have occurred during the COVID-19 pandemic. Similarly, all other being equal, a domestic violence case occurring at a residential location was 22% more likely (than cases at other locations) to have occurred during the pandemic. The pseudo R^2^ was .004, meaning that the model only accounted for .4% of the variation in the dependent variable.

A joint hypothesis test was also conducted to determine whether or not weapon use and resulting arrest jointly contributed to the model fit. The model was run with all variables, and then run again with weapon use and arrest dropped from the model. In comparing the -2 log-likelihood between the two models and obtaining a test chi-square, a significant chi-square of X^2^=8.95, df=2, p=.01 was found, indicating that at least one of the variables tested—either weapon use, resulting arrest, or both—was significant.

### Model 2: Occurrence of domestic violence in shelter-in-place, regressed on multiple variables

The results of the second logistic regression model, predicting occurrence of domestic violence during the shelter-in-place period regressed on several key variables, provided additional significant findings, but again with low explanatory value pseudo R^2^ (Table 4).

In terms of overall model appropriateness, Hosmer-Lemeshow goodness-of-fit test indicated that the logistic response function was appropriate with a non-significant chi-square value of 13.66, p=.62. To check for multicollinearity problems, a VIF test was run, resulting in a low mean VIF of 1.05, indicating no problems. Influential data was assessed with Cook’s distance and was not a problem for this model.

A chi-square test was then performed on the log-likelihood ratio, resulting in a statistically significant overall model (X^2^=27.08, df=5, p<.01). In addition to the significant intercept, in this model, residential location was once again significant. This time, however, resulting arrest was not at all significant, whereas the presence of a child victim was. Odds ratios were then interpreted for residential location (1.638) and child victim (.329). All other being equal, a domestic violence case occurring at a residential location was thus approximately 64% more likely (than cases at other locations) to have occurred during the shelter-in-place. Similarly, a domestic violence case with a child victim was 67% less likely (than cases without child victims) to have occurred during the shelter-in-place order. The pseudo R^2^ was .01, meaning that the model only accounted for 1% of the variation in the dependent variable. This was a slight improvement over the previous model.

A joint hypothesis test was conducted once more to determine whether or not weapon use and resulting arrest jointly contributed to the model fit. This model was run with all variables, and then run again with weapon use and arrest dropped from the model. This time, in comparing the -2-log-likelihood between the two models and obtaining a test chi-square, the chi-square value was not significant at X^2^=1.95, df=2, p=.38, so the null hypothesis cannot be rejected, and the conclusion is that that neither variable was significant. Weapon use and resulting arrest did not jointly contribute to this model, unlike the previous one.

## Conclusions

### Interpretation of Findings

This study offers a rapid comparison of domestic violence cases occurring in a large midwestern city before and during the COVID-19 pandemic, contributing to the lack of literature in this critical area. The two models—one predicting occurrence of domestic violence during the pandemic, and the other predicting occurrence of domestic violence during the shelter-in-place period—offer multidimensionality in analyzing the pandemic’s impact, taking its rapidly-changing nature (both epidemiologically and policy-wise) into account. While both models had low explanatory power (.4% and 1%, respectively), the models were statistically significant at p<.001 (model 1) and p<.01 (model 2). More important, multiple key characteristics (also significant at p<.01 or better) were identified as important predictors of how domestic violence may differ during the pandemic and under shelter-in-place orders.

Residential location was a significant predictor in both models. Location had an even greater impact during the shelter-in-place period, as residential cases were 64% more likely to have occurred during the shelter-in-place order than during the pandemic but before the order. During both periods in 2019 and 2020, cases occurring in a residential location comprised over 75% of all domestic violence cases analyzed, so this study’s finding is not all that notable given the already high numbers of cases occurring in the home. However, this finding also suggests that domestic violence first responders and service providers may need to focus their efforts on residential areas and additional methods of reaching individuals who are staying at home. This finding may also suggest that individuals in this particular area at this particular time were taking precautions and staying at home, which may suggest compliance with shelter-in-place orders.

Two other predictor variables were significant. Arrests were less likely during the pandemic, but only by 20%; this impact did not hold when shelter-in-place was ordered. What did change during shelter-in-place was the presence of child victims—by quite a margin. With cases with child victims being 67% less likely to have occurred during the shelter-in-place order, the question here may be less about occurrence and more about reporting, a point referenced by Piquero and colleagues (2020) in their exploration of domestic violence in Dallas during stay-at-home orders. This finding illuminates a need for further research to understand the relationship between the COVID-19 pandemic and reporting for child victims of domestic violence, particularly around its impacts on mandated reporting numbers and access.

While a minor focus of this analysis, one more notable finding is the total number of reported domestic violence cases during the pandemic as compared to the previous year. With 51.26% (n=2367) of these cases occurring before the pandemic, in this sample, police reports of domestic violence actually went down. This finding contradicts previously reported statistics of domestic violence increases during the COVID-19 pandemic (Bosman, 2020; Leslie & Wilson, 2020), but is corroborated by Mohler and colleagues’ (2020) finding that the overall effect of the pandemic on crime in Los Angeles and Indianapolis was less than expected. It also adds weight to Piquero and colleagues’ (2020) finding of a two-week spike and subsequent drop in domestic violence in Dallas, as well as Bullinger and colleagues’ (2020) estimation of underreporting of domestic violence in Chicago during the period of this study. (For more on crime rates during COVID-19, see also Stickle & Felson, 2020.) While variables and case types may differ across reports, greater attention may be needed on the source of the report—police versus hotline data—than has been available in the U.S., which is an area now being explored by Silverio-Murillo and colleagues (2020) in Mexico City.

### Limitations and Strengths

This study is limited in a few key ways. A major limitation is the lack of available data. Several data sources in Chicago were searched and contacted throughout the research process, but only administrative police data was available at the time of the study. Additionally, this data was limited in the number of available variables, and extensive and rapid data management was needed to ready it for logistic regression analysis. Particularly concerning was the lack of demographic information about perpetrators and victims, which limited the richness of the analysis and the ability to understand the pandemic’s impact on domestic violence for certain communities. It is understandable that data would be sparse during this time, with reduced staffing and competing priorities. Some data, especially confidential domestic violence organization data, is simply not available as quickly as a pandemic would necessitate.

Additionally, there were time and methodological constraints that impacted the depth of analysis with the data available. With more time to explore different crime codes, types of abuse such as telephone harassment, stalking, and order of protection violations could have been included. Finally, low pseudo R^2^ results and the limitation to one metropolitan area reduce generalizability and explanatory power of the findings, significant though they may be.

The study’s strengths are the large sample, the innovation in working with available data, and the rapidness of response. Utilizing a large city’s data allowed for nearly 5,000 domestic violence cases to be analyzed, offering great statistical power for this exploratory study. The study was also able to adapt to available data through a rapid data management strategy that recoded crime codes into dichotomous variables. The use of both pandemic and shelter-in-place timeframes for analysis, with a direct comparison group to the year prior, made for a more robust analysis and provides a glimpse at the rapid changes that data can take on during this unprecedented time. Lastly, the study was able to make quick use of data that was available only a month prior to analysis. Its rapid approach to analysis and writing during such limitations and strain can serve as a model for other social work research during the COVID-19 pandemic.

### Implications

This study provides a glimpse at how domestic violence may differ during the COVID-19 pandemic and its shelter-in-place orders, as well as a few key ways in which those differences manifest. Domestic violence first responders and service providers, as well as researchers, must pivot their approaches to understanding, preventing, and treating this problem given the rapidly-evolving nature of COVID-19 and its impacts on social and economic crisis (Sharma & Borah, 2020). Given the significant findings of cases more likely to occur in residences and less likely to involve child victims during shelter-in-place, additional resources are needed in this critical areas, both to make residential areas safer and to protect children who are vulnerable at this time. The latter point is perhaps the study’s most notable finding, and one in which urgent responses may be needed. With in-person access to mandated reporters like teachers and social workers being restricted due to stay-at-home orders, innovative ways of understanding and responding to children’s experiences in the home are needed to adapt to these new circumstances.

The low explanatory power of the two regression models could be improved with the identification of additional important predictors. For future research to improve, additional sources of data are needed, as are greater availability of variables that might contribute to model explanatory power. Davis and colleagues (2020) made gains in this area by quickly attaching a four-item tool to an actively-running intimate partner violence survey to assess self-reports of COVID-19 and risk of perpetration and victimization. Other information such as the length of wait time for police response, location of first contact with police and service providers, co-occurring protective order violations, and elements of virtual/cyber abuse would be useful to further understanding the problem. Future research could compare domestic violence reported to police, to hotlines, and both during the pandemic, compared to usual reporting rates, to determine which services individuals are accessing during this time that looks so different from the norm. Open data sharing between research and practice realms would assist in this regard.

Lastly, funding must be directed to service providers, to move victims out of dangerous living situations, where more violence is being reported, and into safe spaces. Sheltering in place may create safety from COVID-19, which is of great public benefit. However, if physical and emotional safety are put at greater risk as a result, research and resources are needed to make sure individuals are simultaneously safe from both kinds of threats.

### Note Added

Since the initial submission of this work, global case totals of COVID-19 have gone up by nearly 1000%, and global deaths have more than quadrupled. Given the continued seriousness of the pandemic and need for rapid data analysis, the author is delighted to see research on COVID-19 and domestic violence gain further traction in several since-published articles (Gebrewahd et. al., 2020; Mohler et. al., 2020; Piquero et. al., 2020; Sediri et. al., 2020; Sharma & Borah, 2020; Stickle & Felson, 2020), as well as forthcoming and working papers (Bullinger, Carr, & Packham, 2020; Davis, Gilbar, & Padilla-Medina, 2020; Leslie & Wilson, 2020; Ravindran & Shah, 2020; Silverio-Murillo, Balmori de la Miyar, & Hoehn-Velasco, 2020). The author is indebted to these scholars for their important contributions in the wake of this study.

## Data Availability

All data utilized in this study is available via the Chicago Data Portal. Citation: Chicago Data Portal. (2020). Crimes - 2001 to present [Data file and code book]. Retrieved from https://data.cityofchicago.org/Public-Safety/Crimes-2001-to-present/ijzp-q8t2

## Acknowledgements

The author thanks Brett Drake, Merriah Croston, Maxine Davis, Tonya Edmond, Shenyang Guo, Melissa Jonson-Reid, and Nancy Jacquelyn Perez-Flores for their encouragement and methodological guidance. The author also thanks Amirrah Abou-Youssef, Neha Gill, Lindsay Hawley Nathan, and Andrew Houghton for their on-the-ground wisdom that motivated the continuation of this work, and for their service to the Chicago area community.

